# Humoral SARS-CoV-2 vaccine responses are durable in solid organ transplant recipients with and without HIV

**DOI:** 10.1101/2025.05.07.25327192

**Authors:** Meenakshi M. Rana, Brandy Haydel, Gina Carrara, Charles Gleason, Jacob Mauldin, Komal Srivastava, Sander S Florman, Judith Aberg, Morgan van Kesteren, Jacob Mischka, Juan Manuel Carreño, Gagandeep Singh, Damodara Rao Mendu, TITAN Study Group, Ania Wajnberg, Carlos Cordon-Cardo, Florian Krammer, Viviana Simon

## Abstract

**Background:** Solid organ transplant (SOT) recipients may have a suboptimal humoral immune response to the coronavirus disease 2019 (COVID-19) vaccine, prompting the need for additional doses of vaccine for immunocompromised patients. However, data regarding immune responses to vaccination specifically in SOT recipients with well controlled HIV are lacking.

**Methods:** We conducted a prospective observational cohort single-center study of SOT recipients with and without HIV-1 who had received two doses of mRNA COVID-19 vaccine and were planning to receive additional doses. Severe acute respiratory syndrome coronavirus 2 (SARS-CoV-2) binding and neutralizing antibody responses were measured at several time points after vaccination.

**Findings:** Of the 122 SOT recipients enrolled, 44 (36%) were people with HIV (PWH). Overall, 65% (50/77) of all SOT recipients were seropositive prior to a third vaccine dose. Seropositive SOT recipients with HIV had comparable anti-spike antibody responses at baseline and over time to those without HIV. In addition, HIV status did not impact neutralizing titers in our SOT cohort. Twenty-seven participants were seronegative at baseline; three (11%) were participants with HIV. In addition, 78% (21/27) of participants seroconverted over the duration of the study; of those who remained seronegative, none had HIV, but all were on an antimetabolites.

**Interpretation:** HIV status did not impact longitudinal spike-binding antibody titers or neutralizing titers in SOT recipients.

**Research in context:** 

**Evidence before this study:** Solid organ transplant (SOT) recipients may mount poor humoral immune responses to COVID-19 vaccines, prompting the need for additional vaccine doses in this patient population. Additional risk factors for poor immune response in this population have been described and include for example, age or use of certain immunosuppressant therapies. However, humoral responses to COVID-19 vaccine in SOT recipients with HIV have not previously been described.

**Added value of this study:** We conducted a prospective observational single center study of solid organ transplant recipients with and without HIV and measured SARS-CoV-2 binding and neutralizing antibody responses longitudinally. Our study results demonstrate that HIV status did not appear to be an additional risk factor that affected the durability of spike-antibody titers or neutralizing titers in SOT recipients over time.

**Implications of all the available evidence:** Well-controlled HIV infection is not an additional risk factor in SOT recipients when assessing responses to COVID-19 vaccine. Future studies should continue to focus on other risk factors, such as type of immunosuppressant therapies and timing of vaccination in relationship to transplant.

## Introduction

The advent of the severe acute respiratory syndrome coronavirus 2 (SARS-CoV-2) pandemic in March 2020 had an unprecedented impact on solid organ transplant (SOT) recipients living in New York City. Associated with increased rates of morbidity and mortality in SOT recipients, the looming threat of coronavirus 2019 disease (COVID-19) left many SOT recipients feeling vulnerable. The rapid initial emergency use authorization of the Pfizer BioNTech and then Moderna mRNA vaccines brought hope to this high risk population. However, subsequent early studies soon showed that SOT recipients who received only one or two doses of COVID-19 vaccine had diminished antibody responses to vaccination as compared to others. Boyarsky and colleagues first described this in a SOT cohort of 658 recipients where 46% of recipients did not mount an antibody response despite two doses of vaccine [1]. Subsequent studies have shown improved anti-spike and neutralizing antibody responses following additional vaccine doses in SOT recipients, ultimately leading the US Food and Drug Administration (US FDA) to authorize additional vaccine doses in immunocompromised patients [2–4]. Additional risk factors such as older age, poor graft function, use of mycophenolate and belatacept, and recent use of antithymocyte globulin (ATG) have been associated with poor antibody responses in SOT recipients [5–9].

In contrast to SOT recipients, people with HIV (PWH) who developed COVID-19 appeared to have similar clinical outcomes compared to those without HIV; these findings were more commonly seen in patients on antiretroviral therapy who were virologically suppressed, and severe outcomes due to COVID-19 appeared to correlate more with other co-morbidities [10]. In addition, PWH with suppressed viral loads on antiretroviral therapy (ART) with CD4+ T cells above 200 per mm^3^ appeared to have similar antibody responses after vaccination to those without HIV [11–13]. Despite this, minimal data has been published on SOT recipients with HIV and outcomes related to SARS-CoV-2 infection or vaccination. In one case series, SOT recipients with HIV appeared to have poor clinical outcomes after SARS-CoV-2 infection [14]. However, data regarding immune responses to vaccination specifically in SOT recipients with HIV are lacking.

We conducted a single center, prospective observational study measuring humoral SARS-CoV-2 immune responses in 133 SOT recipients with and without HIV over time following ancestral monovalent COVID-19 vaccination. We hypothesized a third COVID-19 vaccine dose would increase binding antibody titers in all study participants, independent of their HIV status.

## Methods

### Study participants

We conducted a prospective observational cohort single-center study of SOT recipients who had received two doses of mRNA COVID-19 vaccine and were planning to receive additional vaccine doses, henceforth known as the TITAN cohort (**T**racking **I**mmunity through **T**ransplant SARS-CoV-2 **A**ntibodies and **N**eutralization). Solid organ transplant recipients were enrolled into the IRB approved observational research study protocol [STUDY- 16-01215/IRB-16-00971] at the Recanati-Miller Transplant Institute at Mount Sinai Hospital, beginning in August 2021, when the US Centers for Disease Control and Prevention (US CDC) made recommendations for a third vaccine dose as part of the primary series for immunocompromised patients. People with and without HIV-1 as well as varying SARS-CoV-2 immune histories including prior COVID-19 were enrolled between August 2021 and October 2023. All participants with HIV had a suppressed viral load and were on antiretroviral therapy, as part of eligibility criteria for solid organ transplant.

All study participants provided informed consent for participation in research prior to data or sample collection. All human subjects research is reviewed and approved by the Program for the Protection of Human Subjects at the Icahn School of Medicine at Mount Sinai. All clinical research activities and governance of human subjects’ samples are conducted in concordance with US Department of Health and Human Services (DHHS) federal regulations 45 CFR 46. Data regarding vaccination doses, type of SARS-CoV-2 vaccine and SARS-CoV-2 breakthrough infections were collected at baseline and the follow up visits. In addition, basic demographic data including age, gender, race/ethnicity, type of transplant and immunosuppression at the time of each vaccine dose was obtained through the electronic medical record. Baseline blood samples were collected from study participants prior to the third or fourth dose of the ancestral monovalent BNT162b2 (Pfizer) or mRNA-1273 (Moderna) COVID-19 vaccine as well as at multiple time points after, including 30 days, 90 days, 180 days, 300 days, 540 and 720 days after the vaccine dose.

Participants were considered to have completed their participation in the study at the day 300 visit but were asked if they were willing to continue follow-up through the day 720 visit. At each visit, a blood sample was collected, and serum, plasma, and peripheral blood mononuclear cells (PBMCs) were banked for research. Spike-binding IgG antibody levels in sera were measured using either the emergency use-authorized Kantaro SeroKlir or the AdviseDx SARS-CoV-2 IgG II by a Clinical Laboratory Improvement Amendments (CLIA)- certified laboratory. The results of these tests were shared with the study participants.

### SARS-CoV-2 spike binding antibody titers

All sera were analyzed using research grade SARS-CoV-2 spike-binding IgG antibody enzyme-linked immunosorbent assay (ELISA) as described previously [15, 16]. The spike binding endpoint titers and the calculated area under the curve (AUC) provided quantitative antibody titer values with consistency over the entire course of the study.

### Cell lines

African green monkey Vero.E6 cells overexpressing transmembrane protease serine^i^ 2 (TMPRSS2) were cultured in Dulbecco’s modified Eagle medium (DMEM; Thermo Fisher. Scientific) supplemented with 10% fetal bovine serum (FBS), 1x nonessential amino acids (NEAA), 1% penicillin-streptomycin, 3μg of puromycin (InvivoGen) and 100μg of normocin (InvivoGen) at 37°C with 5% CO_2_.

### Primary SARS-CoV-2 isolate

The SARS-CoV-2 isolate USA-WA.1/2020 was used (BEI Resources, NR-52281) since it represents the ancestral SARS-CoV-2 variants that circulated in the beginning of the pandemic (2020/21). Expanded viral stocks of USA-WA.1/2020 were sequence verified and the infectious viral titer (50% tissue culture infectious dose, (TCID_50_)) was determined on Vero.E6-TMPRSS2 cells. All work with infectious SARS-CoV-2 was performed in a biosafety level 3 (BSL-3) facility at the Icahn School of Medicine at Mount Sinai, following standard operating procedures and protocols approved by the Institutional Biosafety Committee.

### SARS-CoV-2 multicycle microneutralization assays

We determined serum-mediated neutralization of SARS-CoV-2 using an established multi-cycle replication assay using SARS-CoV-2 isolates [17]. The day before infection, Vero-E6-TMPRSS2 cells were seeded in 96-well high binding cell culture plates (Costar, #07620009) at a density of 20,000 cells/well in complete Dulbecco’s modified Eagle medium (cDMEM). All sera were heat inactivated at 56 °C for 1 hour. We serially diluted sera over seven points starting at 1:10 in round bottom tissue culture 96 well plates. The initial 1:10 dilutions were followed by three-fold dilutions up to a final dilution of 1:7290 in minimum essential media (MEM; Gibco, #11430-030) supplemented with 2 mM L-glutamine (Gibco, #25030081), 0.1% sodium bicarbonate (w/v, HyClone), 10 mM 4-(2- hydroxyethyl)-1-piperazineethanesulfonic acid (HEPES), 100 U/mL penicillin, 100 μg/mL streptomycin (Gibco) and 0.2% bovine serum albumin (BSA, MP Biomedicals, Cat#810063). Serially diluted remdesivir (Medkoo Bioscience Inc.) was included as an internal control. Diluted sera were incubated with the equivalent of 10,000 TCID50 of WT USA-WA1/2020 for 1 hour at RT. After this incubation, cell culture media was aspirated from Vero-E6-TMPRSS2 plates and 120µL/well of virus-serum mix were gently dispensed down the side of the wells of the cell plates. Infections were left to proceed for 1 hour at 37 °C. Following this incubation period, the inoculum was removed and 100 μL/well of the corresponding sera serial dilutions plus 100 μL/well of infection media supplemented with 2% fetal bovine serum (FBS; Gibco, #10082-147) were added to the cells. Plates were incubated for 48 hours at 37 °C followed by fixation overnight at 4 °C in 200 μL/well of a 10% formaldehyde solution.

For staining of the nucleoprotein, formaldehyde solution was removed, and cells were washed with PBS (pH 7.4) (Gibco) and permeabilized by adding 150 μL/well of 0.1% Triton X- 100 (Fisher Bioreagents) in PBS for 15 min at room temperature (RT). Permeabilization solution was removed, plates were washed with 200 μL/well of PBS (Gibco) twice, and blocked with 3% BSA in PBS for 1 hour at RT. During this time the primary antibody was biotinylated according to manufacturer protocol (Thermo Scientific EZ-Link NHS-PEG4-Biotin). Blocking solution was removed and 100 μL/well of biotinylated mAb 1C7C7, a mouse anti-SARS-CoV-1 nucleoprotein monoclonal antibody generated at the Center for Therapeutic Antibody Development at The Icahn School of Medicine at Mount Sinai ISMMS (Millipore Sigma), at a concentration of 1 μg/mL in 1% BSA in PBS was added for 1 hour at RT. Cells were washed with 200 μL/well of PBS twice and 100 μL/well of horseradish peroxidase (HRP)-conjugated streptavidin (Thermo Fisher Scientific) diluted in 1% BSA in PBS were added at a 1:2000 dilution for 1 hour at RT. Cells were washed twice with PBS, and 100 μL/well of o-phenylenediamine dihydrochloride (Sigmafast OPD; Sigma-Aldrich) were added for 10 min at RT, followed by addition of 50 μL/well of a 3 M HCl solution (Thermo Fisher Scientific). Optical density (OD) was measured (490 nm) using a microplate reader (Synergy H1; Biotek).

After subtraction of background and calculation of the percentage of neutralization with respect to the “virus only” control, a nonlinear regression curve fit analysis was performed to calculate the 50% inhibitory dilution (ID_50_), with top and bottom constraints set to 100% and 0%, respectively for the serially diluted sera.

### SARS-CoV-2 cPass Surrogate Virus Neutralizing Test (sVNT)

This test (GenScript #L00847) detects SARS-CoV-2 antibodies that inhibit the binding of the recombinant receptor binding domain (RBD) of the SARS-CoV-2 spike protein to the human angiotensin converting enzyme 2 receptor. A neutralizing antibody standard curve was established to validate the kit and semi-quantitatively assess neutralizing antibody titers in serially diluted plasma samples. The monoclonal neutralizing antibody (mAb) (GenScript #A02051) was serially diluted with sample dilution buffer (GenScript Ref S1-60) and RBD-HRP solution. This produced a six-point calibration curve with concentrations ranging from 300 U/mL to 4.688 U/mL. The sample dilution buffer was also used for background measurements.

The cPass assay was performed according to the manufacturer’s protocol. Samples and controls were first diluted 1:10 using the dilution buffer, followed by a further 1:2 dilution with the RBD-HRP solution, resulting in a final dilution of 1:20. Pre-diluted samples, standards, and controls were placed into a 96-well sample plate. A preincubation step allowed circulating neutralizing antibodies (nAbs) to bind to the HRP-RBD. After a 30-minute incubation at 37 °C, 100 µl of the diluted samples or controls were added to a 96-well plate pre-coated with recombinant ACE2 protein, followed by another 15-minute incubation at 37 °C. Unbound HRP-RBD or HRP-RBD bound to non-neutralizing antibodies remained on the plate, while the HRP-RBD complexes with circulating nAbs were washed away. After washing, tetramethylbenzidine substrate solution was added, followed by a stop solution that halted the reaction and turned the wells yellow. The optical density (OD) was then measured at 450 nm using a microtiter plate reader. A standard curve of OD450 versus Log Concentration was generated to interpolate the OD450 values from unknown sample dilutions. Samples with high neutralizing antibody concentrations underwent additional serial dilutions to determine end-point titers. Each dilution was tested against the MAB standard curve to calculate the final neutralizing titer for each sample, taking dilution factors into account.

### Statistical Methods

For statistical analysis, SARS-CoV-2 spike binding titers (AUC) and ratios were compared. All statistical tests were run using SciPy (version 1.15.1). Paired samples were tested using the Wilcoxon signed rank test and unpaired samples were tested with the Mann-Whitney U test. Visualizations were created using pandas (2.2.3), NumPy (2.1.3), Matplotlib (3.10.0), and seaborn (0.13.2). Prism 10 software was also used in figure assembly (GraphPad Prism 10.4.1).

## Lead contact

Requests for resources and reagents should be directed to Dr. Viviana Simon

### Materials availability

Reagents used in the study are available upon request to the lead author.

### Data and code availability

No code was used or developed for this study. All data for solid organ transplant recipients will be available from ImmPort under the following identifier: XXX. Data for the PARIS cohort of healthcare workers [18] has been previously published and is available from ImmPort under the identifier SDY2468. Additional information required to reanalyze the data reported in this paper is available upon request.

## Results

Overall, 133 SOT recipients were consented and enrolled into the TITAN cohort study (Supplemental Figure 1). The first 98 participants were enrolled in the 2021-2022 season. Fifteen of these enrollees were participants with HIV. To better evaluate responses between SOT recipients with and without HIV, additional participants were enrolled in the 2022-2023 and 2023-2024 seasons. Thirteen SOT recipients with HIV were enrolled in the 2022-2023 season and 22 were enrolled in the 2023-2024 season. 122 participants were included in the initial cohort, with 77 participants having both a pre- and post-third dose vaccine sample. In addition, 54% (66/122) of participants received additional vaccine doses after completing the 3-dose primary immunization series. Of these, 66 participants received a total of four vaccine doses, 29 received a total of five doses and nine had a total of six immunizations (Table 1). Eleven participants were excluded from all analysis due to study withdrawal (N: 4), incomplete primary immunization series (N: 1), or lack of banked biospecimen (N: 6, see also Supplemental Figure 1).

**TABLE 1:** Demographics and cohort description

| Demographics |  |  |  |
| --- | --- | --- | --- |
| HIV Status | HIV+ (n=44) | HIV- (n=78) | Total (n=122) |
| Gender |  |  |  |
| Female | 14 (32%) | 43 (55%) | 57 (47%) |
| Male | 30 (68%) | 35 (45%) | 65 (53%) |
| Race |  |  |  |
| Asian | 2 (5%) | 7 (9%) | 9 (7%) |
| Black or African American | 21 (48%) | 15 (19%) | 36 (30%) |
| White | 8 (18%) | 14 (19%) | 22 (18%) |
| Other/Unspecified | 13 (29%) | 42 (11%) | 55 (45%) |
| Ethnicity |  |  |  |
| Hispanic or Latino | 15 (34%) | 37 (47%) | 52 (43%) |
| Transplant Type |  |  |  |
| Kidney | 34 (77%) | 46 (59%) | 80 (66%) |
| Liver | 7 (16%) | 25 (32%) | 32 (26%) |
| Multi | 2 (5%) | 6 (8%) | 8 (7%) |
| Other (Trachea) | 0 (0%) | 1 (1%) | 1 (1%) |
| Transplant Timing |  |  |  |
| Transplant Prior to Immunization | 15 (34%) | 65 (83%) | 80 (66%) |
| Transplant Between Dose 1 and Dose 3 | 11 (25%) | 13 (17%) | 24 (20%) |
| Transplant After Dose 3 | 18 (41%) | 0 (0%) | 18 (15%) |
| COVID Pre-Enrollment? |  |  |  |
| Yes | 9 (30%) | 14 (18%) | 23 (21%) |
| Primary Vaccine Type |  |  |  |
| Moderna | 14 (35%) | 22 (30%) | 36 (30%) |
| Pfizer | 30 (65%) | 56 (70%) | 86 (70%) |
| Additional Vaccine Doses |  |  |  |
| Dose 4 | 33 (75%) | 33 (45%) | 66 (54%) |
| Dose 5 | 21 (47%) | 8 (11%) | 29 (23%) |
| Dose 6 | 9 (20%) | 0 (0%) | 9 (7%) |
| Medications at Third Dose |  |  |  |
| CNI | 2 (4%) | 12 (16%) | 14 (11%) |
| CNI+Pred | 1 (2%) | 6 (8%) | 7 (7%) |
| Other | 2 (4%) | 2 (3%) | 4 (3%) |
| CNI, with AM | 6 (14%) | 20 (26%) | 26 (21%) |
| CNI+Pred, with AM | 14 (32%) | 36 (46%) | 50 (42%) |
| Other, with AM | 1 (2%) | 2 (3%) | 3 (2%) |

Other demographics stratified by HIV status are described in Table 1. Of the 122 SOT recipients included in this analysis, 44 (36%) were PWH. While 47% of the overall TITAN cohort were women, men comprised most of the cohort with HIV (30/44, 68%). In addition, 48% of the HIV SOT cohort identified as Black and 34% identified as Hispanic, of those without HIV, 19% of SOT recipients identified as Black and 47% identified as Hispanic. Viral replication in PWH was well controlled on antiretroviral combination therapy: 93% (41/44) of participants had viral load values below detection (<50 copies). All PWH had viral load values under 200 copies within six months of vaccine dose 3. The median absolute CD4 count of the HIV positive SOT was 388 (range 64-1216). While 66% of the overall TITAN cohort received their vaccine series after transplant, most of those with HIV had two doses of vaccine prior to solid organ transplantation (61%).

Seropositive SOT recipients with HIV had comparable baseline anti-spike antibody responses to those without HIV. Both groups received additional doses of vaccine and had episodes of COVID-19 but maintained comparable antibody responses throughout the follow-up period (Figure 1). Transplant recipients overall had lower spike-binding antibody titers both before and after the third vaccine dose (p<0.001, Mann-Whitney U) when compared to a cohort of 190 healthcare workers (Figure 2A, [18]). However, SOT recipients with and without HIV both had increases in the geometric mean titer thirty days after the 3^rd^ dose (Figure 2B). Of the 27 participants who were seronegative at baseline, the third dose of vaccine resulted in seroconversion for 1/3 participants with HIV and for 8/24 participants without HIV (Figure 2B).

**Figure 1:**
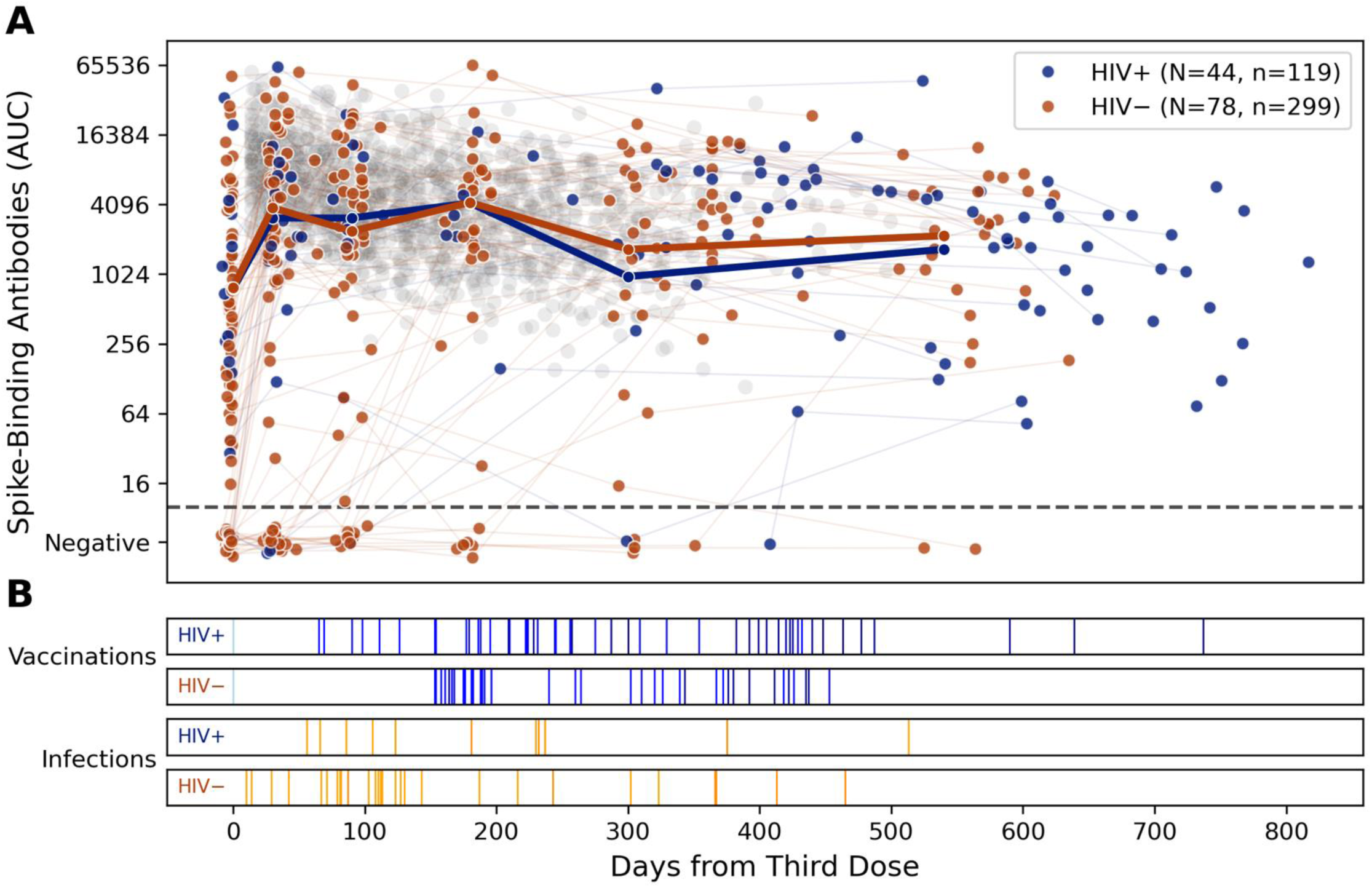
Longitudinal SARS-CoV-2 spike-binding antibody titers in TITAN show comparable responses in solid organ transplant recipients with and without HIV. Each point represents a study visit where spike binding antibody titers (AUC) were measured (A). Visits are colored by the HIV status of the participant. Visits are graphed relative to days from an additional (third) dose of the COVID-19 vaccine. A geometric mean titer for seropositive samples for each classification is plotted over the sample set. Samples testing negative for spike binding antibodies are slightly jittered to prevent overlap and preserve visual clarity. In grey, spike-binding antibodies measured in 1,326 samples from 223 healthy control participants collected after a third mRNA vaccine dose are plotted as a reference. Below (B), vaccination (blue) and infection (orange) events in the transplant recipients are graphed in raster plots separated by HIV infection status.

**Figure 2:**
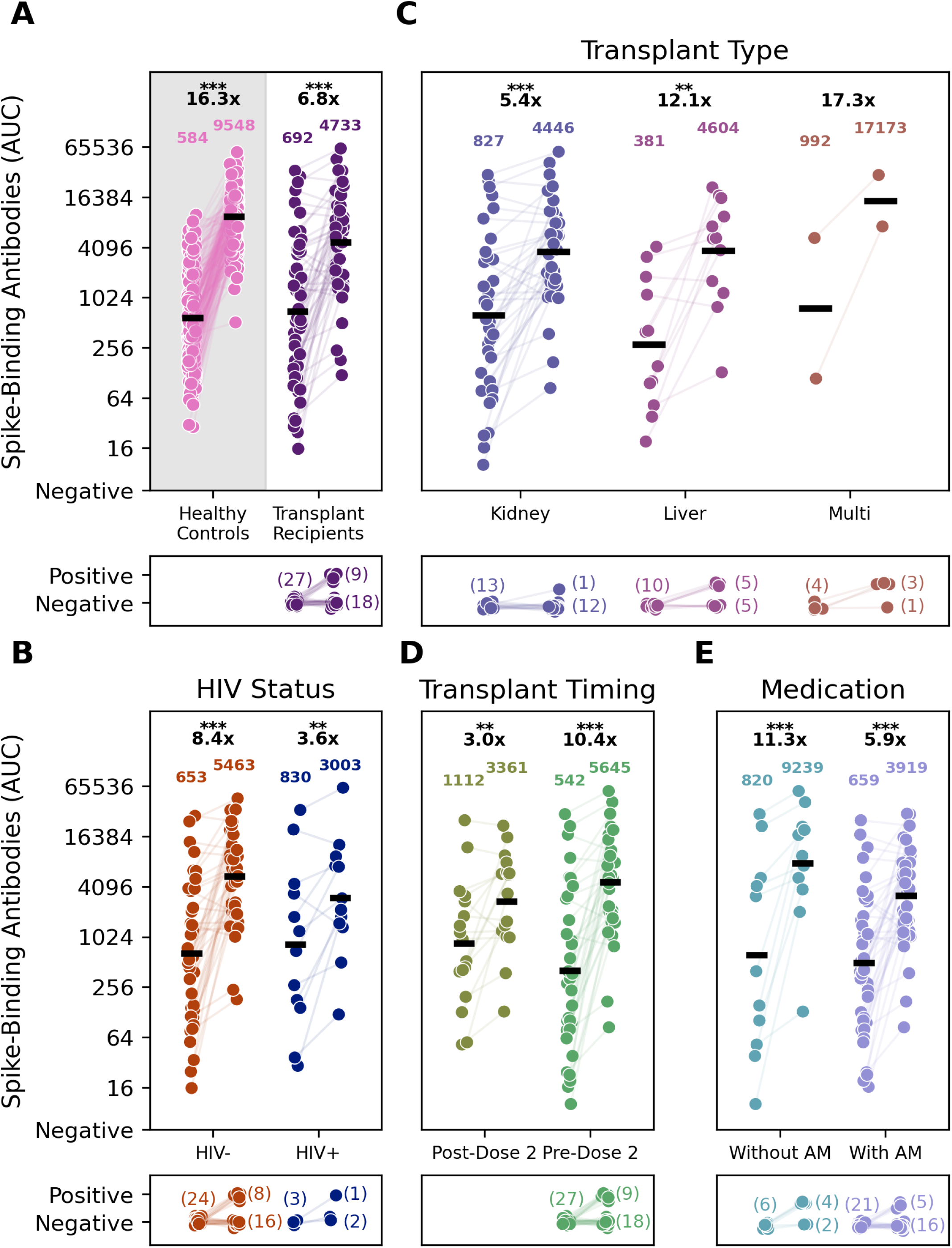
Transplant timing, transplant type, and medication at time of vaccination affect solid organ transplant recipients’ (SOTR) peak responses to an additional dose of the vaccine. Pre-third dose and 30 days post-third dose timepoints are plotted for 77 participants with in- window visits for both timepoints (n=154). The geometric mean titer for the samples is shown above the timepoint. The fold change from baseline to the second timepoint in seropositive samples at baseline is shown above the geometric mean titer. Two and three asterisks above the fold change indicate a p-value of <0.01 and <0.001, respectively. Seroconversions in each group shown are indicated by the graphs below each plot. A: Comparison of Spike binding antibody titers of 190 PARIS participants (healthy controls) and TITAN participants (Transplant recipients) before and after receiving a third COVID-19 vaccine dose. The bottom panel shows the number of participants who sero-converted upon vaccination. B: Spike binding antibody titers measured before and after the third COVID-19 vaccine dose are plotted by HIV serostatus (HIV positive dark blue N=15; HIV negative dark orange; N=62). C: Spike binding antibody titers measured before and after the third COVID-19 vaccine dose are plotted by the type of solid organ transplant received. Kidney SOT recipients are in violet (N=49), Liver SOTRs are light purple (N=22), and Multiple SOTRs are in light red (N=6). D: Spike binding antibody titers measured before and after the third COVID-19 vaccine dose are plotted depending on when participants received the transplant relative to the third vaccine dose. Participants who received their third dose after their transplant are in olive (N=17), and participants who received their transplant before are in green (N=60). E: Spike binding antibody titers measured before and after the third COVID-19 vaccine dose are plotted depending on whether antimetabolites (AM) were a part of their immunosuppressive medication at the time of vaccination. Values for participants taking AM (N=52) are plotted in lavender and those from participants not taking AM (N=25) are in teal.

Kidney transplant recipients made up 66% of the full cohort and 77% of participants with HIV. Seropositive kidney and liver transplant recipients all had an increase in their geometric mean titers after an additional third dose of vaccine (Figure 2C). Overall, only 1/12 kidney transplant recipients seroconverted with the third vaccine dose but 5/10 liver transplant recipients and 3/4 multiorgan transplant recipients seroconverted (Figure 2C). All participants who received the first two doses of a vaccine series before transplant were seropositive at baseline, prior to third dose. All seronegative SOT recipients (27/77, 35%) were vaccinated after transplant. However, even seropositive participants vaccinated after transplant demonstrated a robust response to the third dose, with a 10.4-fold change in their geometric mean titer (Figure 2D).

More SOT recipients on antimetabolites were seronegative at baseline (21/52, 40%, vs 6/25, 24%). Four out of six (67%) SOT recipients who were not on an antimetabolites seroconverted after the third additional dose, in comparison to five out of 21 (24%) SOT recipients on an antimetabolite (Figure 2E). In addition to the 9/27 participants who seroconverted after the third dose, 12 participants seroconverted on study following SARS-CoV-2infection (8/12), a fourth vaccine dose (1/12), both a fourth vaccine dose and SARS-CoV- 2 infection (1/12) and to suspected SARS-CoV-2 exposures that were either not reported or asymptomatic infection (2/12). Over the full duration of the study, ultimately 21 participants seroconverted while six did not. Of those who remained seronegative, none had HIV, and all were on an antimetabolites—the majority of these were kidney transplant recipients (Figure 3).

**Figure 3:**
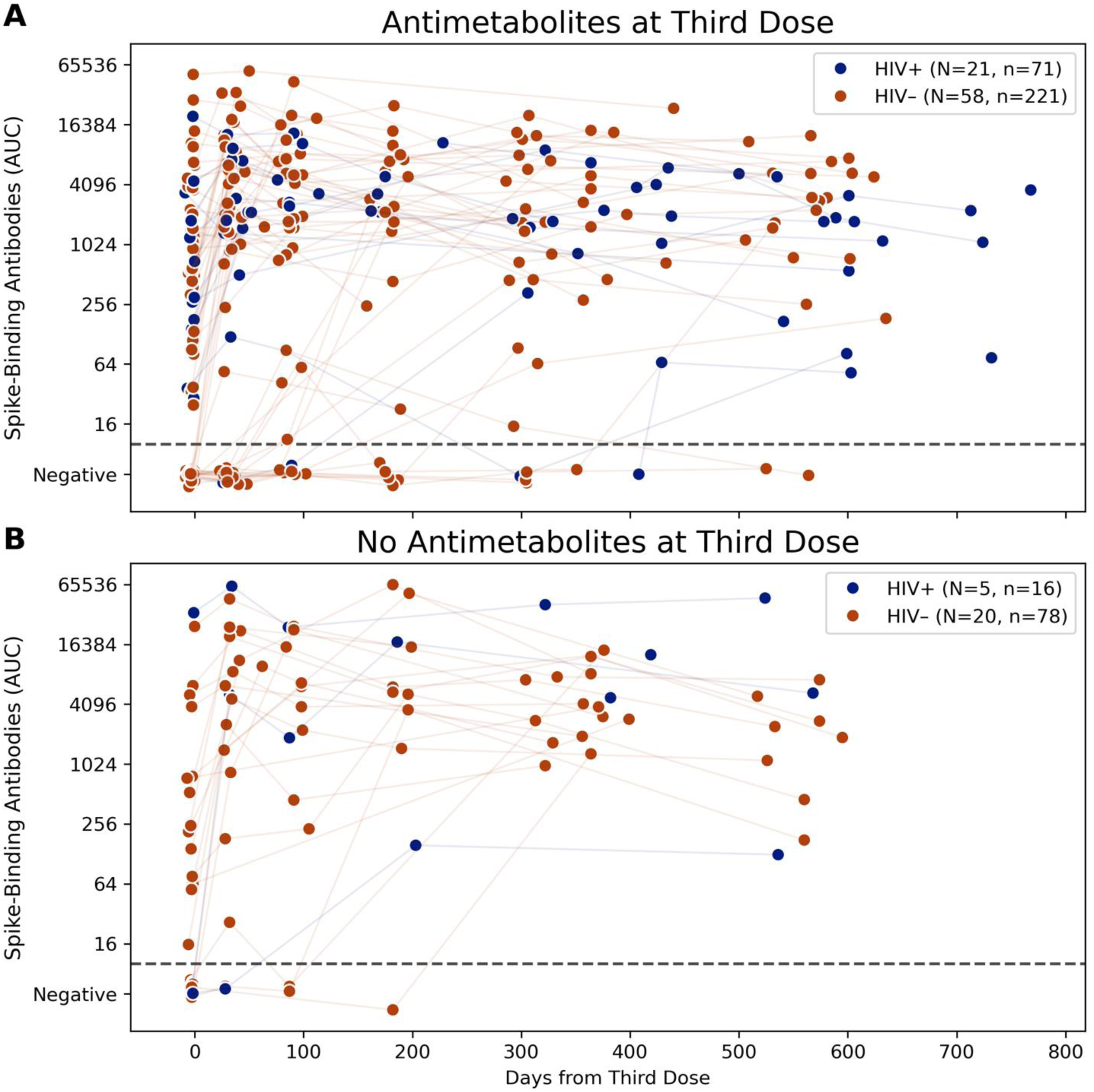
Antimetabolites negatively influence humoral SARS-CoV-2 responses. Longitudinal spike-binding antibody titers measured in solid organ transplant recipients whose medications at time of third dose included antimetabolites (A, 79 participants) and not (B, 25 participants). Participants are classified by HIV status with HIV+ SOTRs in dark blue and HIV- SOTRs in dark orange. The connecting line identifies antibody values from the same study participant. Participants who were not on immunosuppressive medications at the time of their third dose but joined the study with a qualifying booster dose were excluded.

**Figure 4:**
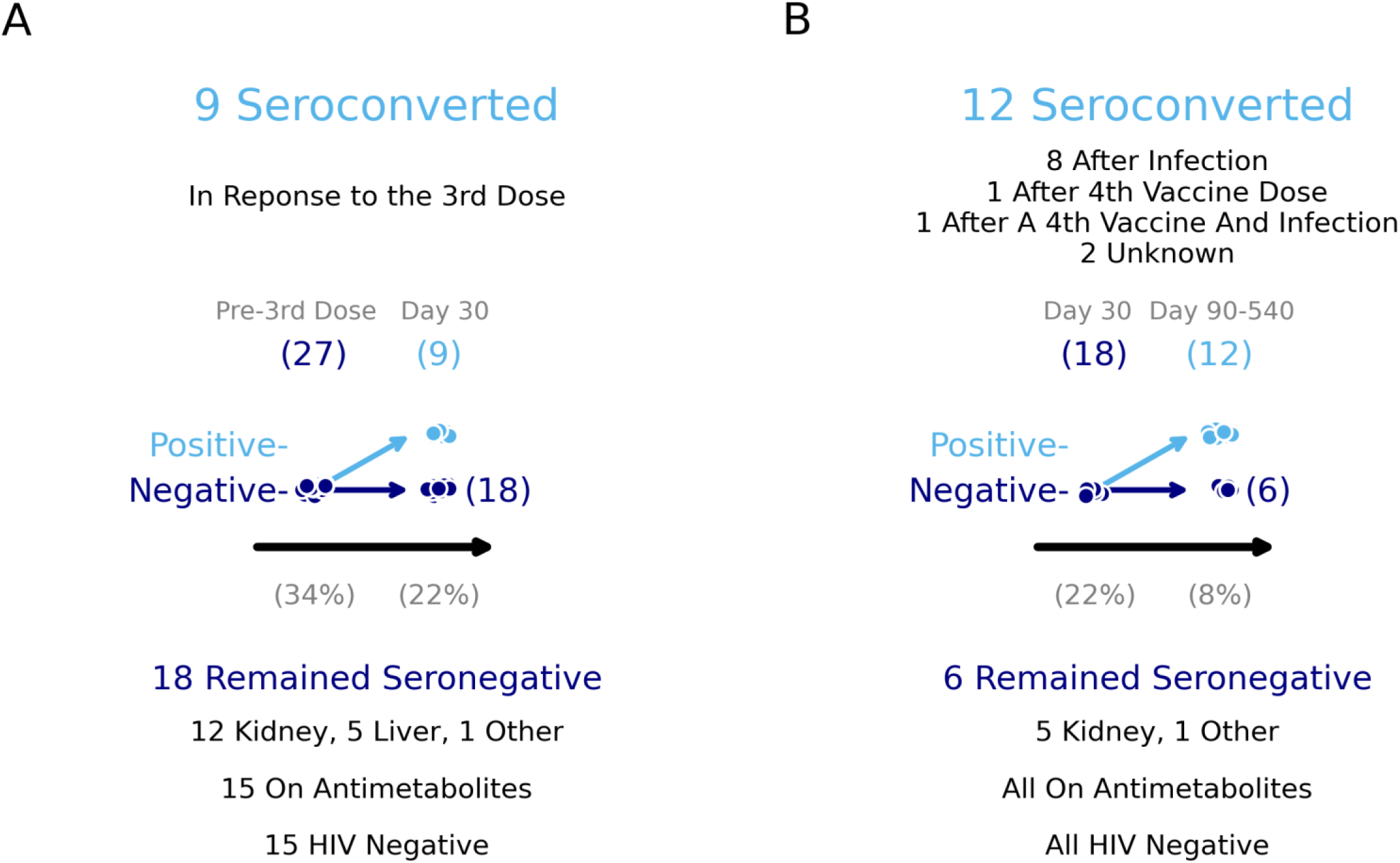
Identification of the variables associated with SARS-CoV-2 seroconversion in response to the third vaccine dose. Seronegative participants who seroconverted during the time course of TITAN did so in response to several events in addition to a third dose of the COVID-19 vaccine. Pre-3rd dose and post-3rd dose timepoints for participants who were seronegative at baseline are shown. Participants who stayed seronegative were plotted again, and further seroconversions over the remaining time course of the study are shown. The seronegative participants as a percentage of the TITAN cohort are shown below the x-axis in both plots. A: The number of participants who were seronegative before the third vaccine dose are plotted. Participants who mounted spike binding antibodies above the limit of detection within 30 days of vaccination were categorized as having seroconverted (n=9). Details on the 18 participants who remained seronegative are below the plot. B: The number of participants who remained seronegative 30 days after the third dose are plotted. Participants who seroconverted at or between 90 to 540 days post-3rd dose are moved to positive. Contextual information on the seroconversion can be found above the plot. Further details on the remaining seronegative participants can be found below the plot.

To complement our serological analysis, we sub-selected participants with longitudinal follow-up to test for neutralization against ancestral SARS-CoV-2 to match the strain included in participants’ third and fourth vaccine doses. To measure neutralization activity, we used both a multi-cycle microneutralization assay with replication-competent SARS-CoV-2 isolates as well as an ELISA measuring inhibition of ancestral SARS-CoV-2 RBD to ACE2 (cPass). Twenty-four patients had neutralizing titers measured over the duration of the study. All participants with and without HIV had an increase in these titers measured against replication-competent ancestral SARS-CoV-2 after vaccine doses 3 and 4 that correlated well with an increase in anti-spike antibody measurements (Figure 5). Results from this multi-cycle microneutralization assay were in concordance (Pearson r=0.91) with those from the cPass assay. The greater dynamic range of cPass also captures longitudinal changes in one participant with sustained high neutralizing titers throughout the study (Figure 5A).

**Figure 5:**
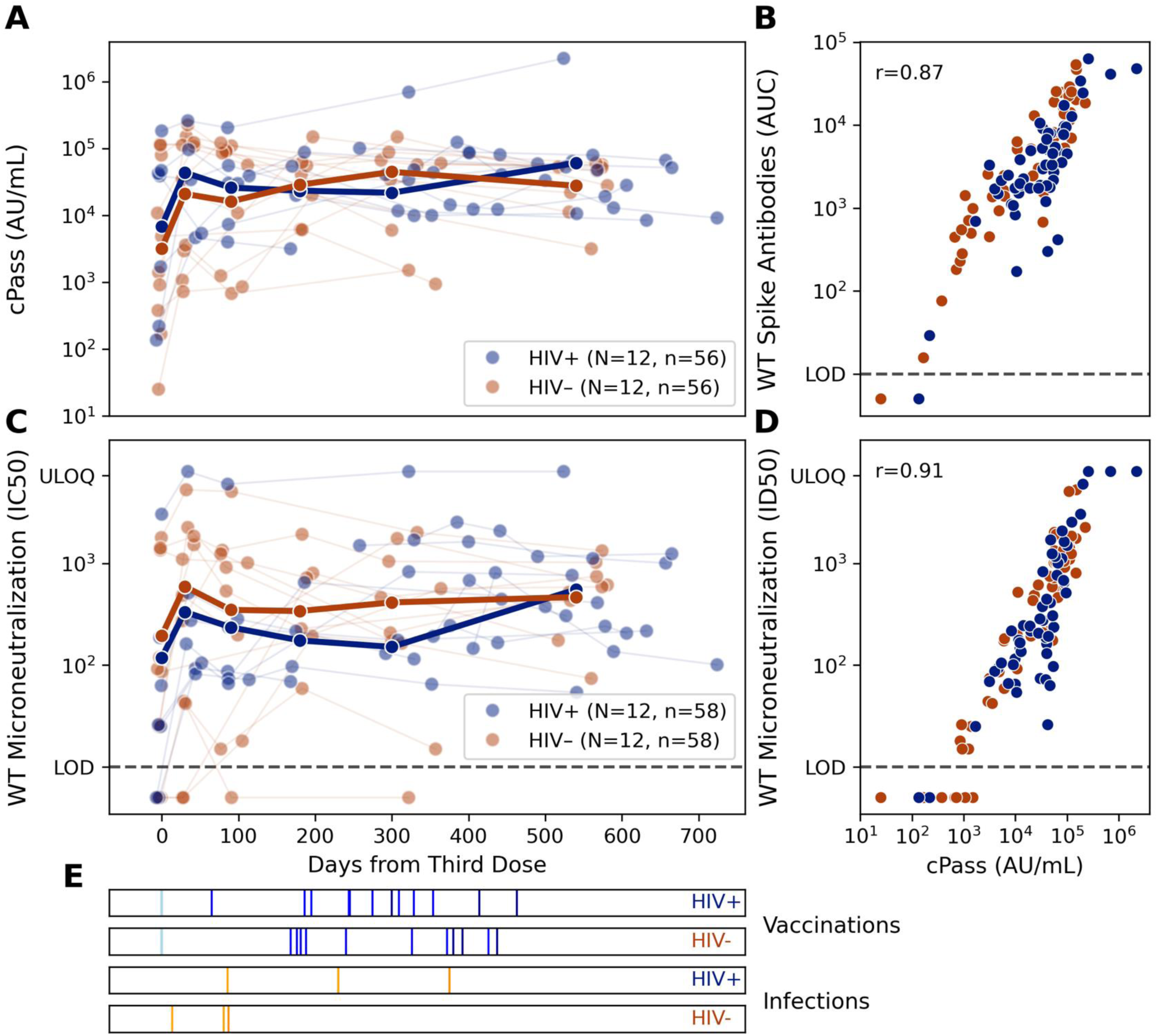
Virus neutralization activity is maintained over prolonged duration. A: The virus neutralization activity of longitudinal sera from 12 HIV negative and 12 HIV positive study participants was determined using surrogate test evaluating RBD ACE2 binding inhibition (A) or a multi-cycle microneutralization assays with replication competent SARS-CoV-2 (C). Vaccinations (blue) and infections (orange) for the selected participants are shown in raster plots below the longitudinal neutralization plots. B: Values for cPass surrogate neutralization are plotted against matched spike-binding antibody titers D: Values for cPass surrogate neutralization are plotted against matched multi-cycle microneutralization infectious dilution 50 values. E: Raster plots indicating SARS-CoV-2 vaccination and infection events in participants included in the longitudinal series are plotted below panels A and C, separated by HIV status.

## Discussion

Our findings demonstrate that SOT recipients with and without HIV have comparable longitudinal anti-spike antibody responses over time. Overall, 65% of SOT recipients were seropositive prior to the third vaccine dose. While this may be higher than what has been previously been reported in other SOT studies, we enrolled patients with and without pre-existing immunity due to COVID-19 infection which may have impacted baseline antibody titers [1, 3]. In addition, we included participants who were vaccinated with a two-dose series prior to transplant, which also would explain higher baseline seropositivity rates when compared to studies that only included SOT recipients who were vaccinated after transplant [19].

SOT recipients, irrespective of HIV status, transplant type, timing or antimetabolite use, appeared to have an increase in their geometric mean SARS-CoV-2 spike binding antibody titers after a third dose of the COVID-19 vaccine. In addition, a third vaccine dose resulted in seroconversion for nine study participants. Over time, twenty-one participants seroconverted, either because of additional vaccine doses or SARS-CoV-2 infection. Similarly, in the MELODY study, a higher number of vaccine doses (five versus three) or previous COVID-19 infection were more likely to correlate with a positive antibody response in 9927 SOT recipients [19]. This stands in contrast to healthy healthcare workers, enrolled in the PARIS study, who were followed longitudinally with pre- and post- primary and booster vaccine series [18]. In this study, all healthy participants had significant increases in their spike binding antibody titers after only the second dose of vaccine. In addition, our data suggest that participants enrolled in the PARIS cohort had significantly higher spike-binding antibody titers before and after the third vaccine dose when compared to SOT recipients. Our data corroborates previous findings that suggest that SOT recipients have poorer antibody responses and appear to benefit from a third dose of vaccine, and that even after a third dose of the vaccine, SOT recipients who remain vulnerable benefit from further doses.

We found that when comparing seropositive vs seronegative SOT recipients, comparable numbers of SOT recipients with and without HIV were found in each cohort. In addition, virus neutralization activity was measured longitudinally and remained durable throughout the study period against the ancestral SARS-CoV-2 variant. These neutralizing antibody responses correlated with spike binding antibody responses and appeared independent of HIV status in SOT recipients.

While HIV status did not appear to impact the humoral immune response in our SOT cohort, we did find that two other factors—the use of antimetabolite immunosuppressive therapy and timing of vaccination—appeared to impact anti-spike antibody titers. First, we found that more SOT recipients on an antimetabolite as part of their immunosuppressive regimens were seronegative at baseline than those not on an antimetabolite. These findings have been previously reported and at least two meta-analyses have shown that antimetabolite use has correlated with poor antibody response [8, 20]. In fact, higher trough mycophenolate concentrations have also been shown to correlate with lower antibody titers, supporting a dose-dependent effect [21]. In addition, our study demonstrates the long-term impact of antimetabolites on antibody response; all participants who did not seroconvert were on an antimetabolite. Our findings support the premise behind other studies currently looking at holding anti-metabolites at the time of vaccination to assess if higher antibody levels can be achieved without compromising graft function. In fact, Schrezenmeir and colleagues have shown that temporary withdrawal of mycophenolate for five weeks in transplant recipients resulted in seroconversion after a fourth dose in 76% of patients at day 32 after vaccine [22]. Other prospective randomized studies, such as the COVID Protection after Transplant- Immunosuppression Reduction (CPAT-ISR) are ongoing to assess whether this is a feasible approach in SOT recipients (ClinicalTrials.gov, NCT05077254).

In addition, we found that all SOT candidates who received a primary two dose vaccine series prior to transplant were seropositive. Despite this, seropositive SOT participants also benefited from an additional dose, with a 6.8-fold change in geometric mean titer (Figure 2A) and a 10.4-fold change in geometric mean titer if vaccinated after transplant (Figure 2D). However, all seronegative participants were those who were vaccinated after transplant. These findings highlight the importance of emphasizing vaccination to candidates prior to transplantation. Previous studies have shown that patients are more likely to mount an immune response to vaccines, for example the influenza vaccine, before transplant, supporting guidelines that recommend vaccination prior to transplantation [23, 24]. Our data suggest similar findings for COVID-19 vaccination, supporting recommendations that whenever possible transplant candidates should receive at least a primary series before transplant. Grupper *et al.* demonstrated similar findings for a cohort of 19 kidney transplant recipients vaccinated before transplant, demonstrating higher geometric mean titers in this cohort as compared to those vaccinated after transplant [25]. Our study demonstrates similar findings, but with a larger cohort of liver, kidney and multiorgan transplants including participants with HIV.

Lastly, we had several SOT recipients who remained seronegative during the follow up study despite attempts at revaccination. In these situations, other targeted interventions should be considered, such as the use of antibody formulations for COVID-19 pre-exposure prophylaxis [26]. While not routinely recommended for clinical use, anti-spike antibody assays in SOT recipients can be used to help identify patients for whom such therapies may be helpful [27].

Limitations of our study include that we could not assess the impact of vaccines only over time on antibody responses, as we enrolled participants with varying SARS-CoV-2 immune histories. However, our study mimicked a more “real-world” scenario that included all patients with breakthrough SARS-CoV-2 infections and receipt of vaccines at different time points before and after transplant. Another limitation was the smaller sample size of our cohort, which precluded more detailed subgroup statistical analysis. Despite this, we had a large sample size of SOT recipients with HIV, and were able to follow this cohort over time, collecting multiple time points of spike-binding antibody responses. Other SOT studies looking at SARS-CoV-2 immune responses have not specifically looked at SOT recipients with HIV.

In conclusion, HIV status did not impact longitudinal spike-binding antibody titers or neutralizing titers in SOT recipients. However, timing of transplant in relationship to vaccination and the use of antimetabolites as part of immunosuppressive regimen may be more clinically relevant when looking at immune responses to SARS-CoV-2 in SOT recipients. Future prospective studies should focus on SOT recipients with these specific risk factors; in addition, anti-spike antibody assays may be a helpful tool in identifying higher risk patients who may benefit from additional vaccine doses.

### Study group/Consortia

The members of the TITAN study group in alphabetical order are: Ashley Aracena, Harm van Bakel, Yuexing Chen, Ana Gonzalez-Reiche, Giulio Kleiner, Neko Lyttle, Brian C. Monahan, Jessica Nardulli, Emilia Mia Sordillo, Reima Ramsamooj.

### Data sharing statement

No code was used or developed for this study. Serology data for solid organ transplant recipients is available from ImmPort under the identifier (will be added once submission is completed). Data for the PARIS cohort of healthcare workers [18] has been previously published and is available from ImmPort under the identifier SDY2468.

## Data Availability

All data produced in the present study are available upon reasonable request to the authors

## Acknowledgments

This research would not have been possible without the long-term support of our generous study participants. This work was supported in part through the computational and data resources and staff expertise provided by Scientific Computing and Data at the Icahn School of Medicine at Mount Sinai and supported by the Clinical and Translational Science Award (CTSA) grant UL1TR004419 from the National Center for Advancing Translational Sciences. We are also thankful to the Mount Sinai Pathogen Surveillance Program for providing expert sequencing of representative viral isolates.

This effort was supported by the Serological Sciences Network (SeroNet) Capacity Building Center Task Order 21X092F1 in part with Federal funds from the National Cancer Institute, National Institutes of Health, under Contract No. 75N91019D00024, Task Order No. 75N91021F00001. The content of this publication does not necessarily reflect the views or policies of the Department of Health and Human Services, nor does mention of trade names, commercial products or organizations imply endorsement by the U.S. Government.

The Conventional Biocontainment Facility (CBF) is a NIH BSL3/BSL3 facility that is part of the BSL-3 Biocontainment CoRE. This Core is supported by subsidies from the ISMMS Dean’s Office and by investigator support through a cost recovery mechanism. Research reported in this publication was supported by the National Institute of Allergy and Infectious Diseases of the National Institutes of Health under Award Number G20AI174733 (Dr. R.A. Albrecht). The content is solely the responsibility of the authors and does not necessarily represent the official views of the National Institutes of Health.

## Contributors

MR, SF, JA, AW, CCC, FK, VS were responsible for the conception and design of the study. MR, GC and BH were responsible for enrollment of patients and collection of samples. JA, SF, AW, provided clinical evaluations. JMC, CG, JM, JM, GS, MvK, JM, DRM, KS were responsible for data generation, sample analysis and data analysis. CCC, VS, FK and KS raised financial support. The manuscript was written by MR, CG, JM, KS and VS. All authors reviewed and approved the manuscript.

---

**Supplemental Figure 1:**
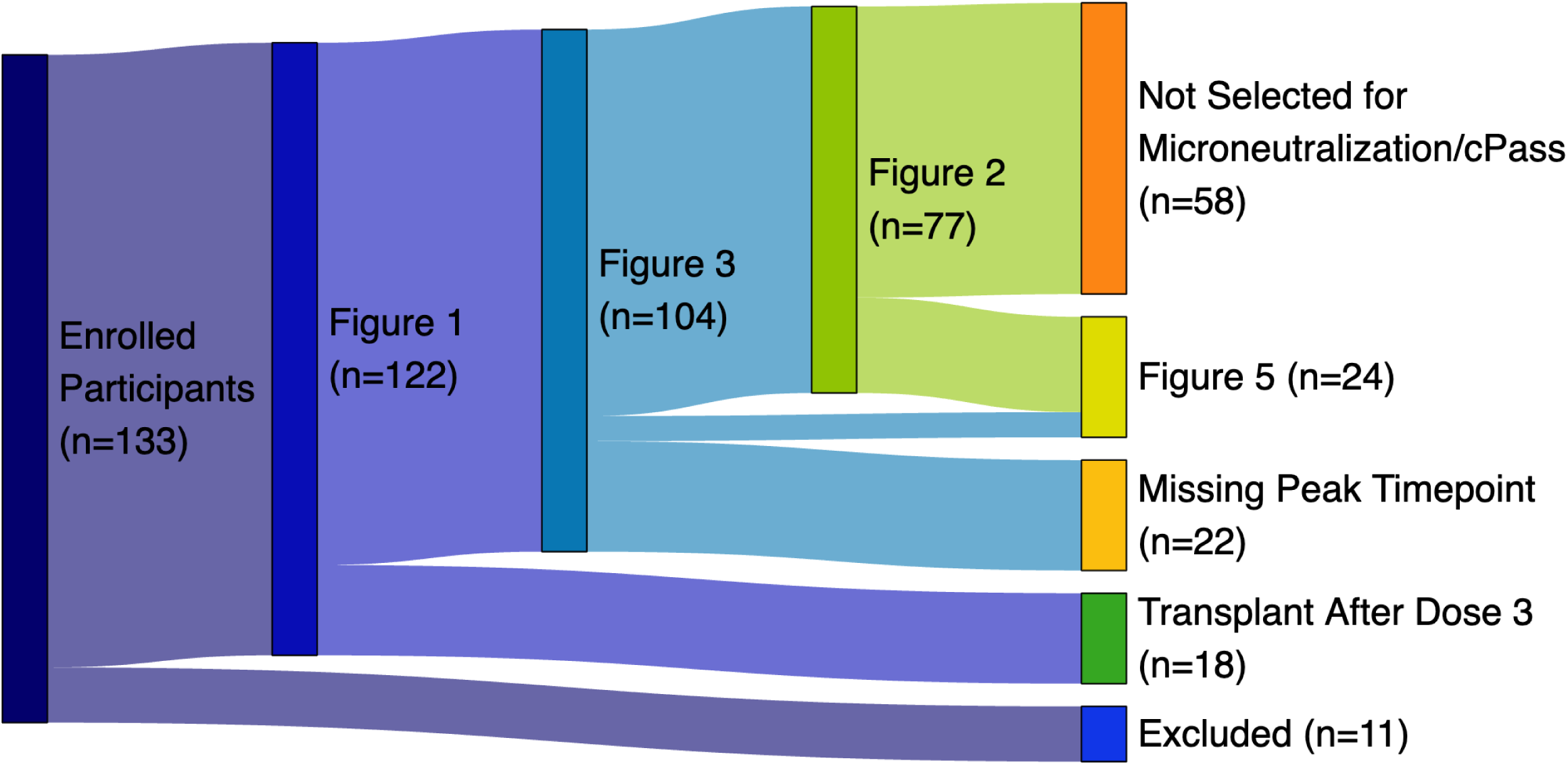
Summary of the TITAN study participants data included in the different experiments This Sankey diagram illustrates the selection and classification of participants through various figures in the paper. The initial pool consists of 133 participants in the TITAN study. These participants were subsequently filtered into different figures based on various criteria. All active or completed participants are shown in Figure 1. Participants with transplant prior to vaccine dose 3 are shown in Figure 3, and of those participants with a pre-dose 3 and peak post-dose 3 timepoint are included in Figure 2. For Figure 5, 24 participants with 4-5 longitudinal follow-up timepoints were selected for additional experiments measuring microneutralization and RBD ACE2 binding inhibition (cPass).

